# A Preoperative Estimate of Central Venous Pressure Is Associated with Early Fontan Failure

**DOI:** 10.1101/2019.12.28.19016048

**Authors:** Michael A. Quail, Ignatius Chan, Shiv Sarna, Marina Hughes, Vivek Muthurangu

**Affiliations:** Centre for Cardiovascular Imaging, UCL Institute of Cardiovascular Science & Great Ormond Street Hospital for Children, London, UK; Adult Congenital Heart Disease Unit, Royal Papworth Hospital NHS Foundation Trust, Papworth Road, Cambridge Biomedical Campus, Cambridge, UK

**Author notes:** **Correspondence to:** Dr Michael Quail, Level 3 ZCR, Great Ormond Street Hospital for Children, Great Ormond Street, London, WC1N 3JH.

## Abstract

**Objective:** Early Fontan Failure (EFF) is a serious complication following total cavopulmonary connection (TCPC), characterised by high central venous pressure (CVP), low cardiac output and resistance to medical therapy. This study aimed to estimate post-operative CVP in TCPC patients (CVP_TCPC_) using data routinely collected during pre-operative assessment. We sought to determine if this metric correlated with measured post-operative CVP and if it was associated with EFF.

**Methods:** In this retrospective study, CVP_TCPC_ was estimated in 131 patients undergoing pre-TCPC assessment by cardiac magnetic resonance imaging and CVP measurement under general anaesthesia. Post-operative CVP during the first 24hours in ICU was collected from electronic patient records in a subset of patients. EFF was defined as death, transplantation, TCPC takedown or emergency fenestration within the first 30days.

**Results:** Estimated CVP_TCPC_ correlated significantly with CVP_ICU_ (r=0.26, p=0.03), particularly in patients without a fenestration (r=0.45, p=0.01). CVP_TCPC_ was significantly associated with EFF (Odds Ratio [OR] 1.12 (1.02-1.24), p=0.02). A threshold of CVP_TCPC_ ≥33mmHg was found to have the highest specificity (90%) and sensitivity (67%) for identifying EFF (area under receiver operating curve, AUC = 0.79), OR 18.8 (3.1-114), p=0.001. This association was stronger in patients with single SVCs.

**Conclusions:** Estimated CVP_TCPC_ is an easily calculated metric combining pre-operative pressure and flow data. Higher CVP_TCPC_ is associated with an increased risk of EFF and is correlated with directly measured post-TCPC pressure. Identification of patients at risk of EFF has the potential to guide risk mitigation strategies.

## Introduction

Early Fontan failure (EFF) is a malignant haemodynamic state which occurs in the early post-operative period following total-cavopulmonary connection (TCPC). EFF is primarily characterised by high central venous pressure (CVP), as well as low cardiac output and resistance to medical therapy. Importantly, EFF may result in death, take-down of the TCPC, emergency fenestration or cardiac transplantation. ^1 2^

It is recognized that mean CVP rises linearly with both pulmonary blood flow (Q_p_) and pulmonary vascular resistance (PVR) in patients with cavo-pulmonary connections. Thus, the transition from the bidirectional cavopulmonary connection (BCPC) to the TCPC must result in increased CVP, due to the increase in Q_p_. Patients who experience large rises in CVP may be at increased risk of EFF.

Unfortunately, pre-operative biomarkers for EFF are lacking. ^3 4^ Given the pathophysiology of EFF, identification of a postoperative high CVP phenotype would be desirable to both to inform surgical risk and guide mitigation strategies (e.g. elective fenestration).

One possibility is to use pressure and flow data, routinely acquired in the pre-operative BCPC state, to derive an estimate of CVP following TCPC completion.

In this study we aimed to i) estimate CVP in the immediate TCPC post-operative period using data routinely collected during pre-operative cardiovascular magnetic resonance (CMR) and ii) determine the association, if any, with CVP measured in ICU and iii) assess if metrics were associated with EFF.

## Methods

### Study population

The study cohort included all children between April 2005 and September 2017 who underwent elective pre-TCPC CMR assessment in whom a complete CMR flow and CVP dataset were available:131 patients from a total population of 147. Demographic and clinical details were obtained from the medical record.

Informed consent for the use of imaging data was obtained from all parents or guardians of the patients included in this study. The study protocol conforms to the ethical guidelines of the 1975 Declaration of Helsinki and was approved by the local committee of the UK national research ethics service (06/Q0508/124).

### CMR protocol

All CMR studies were undertaken on a 1.5 T MR scanner (Avanto; Siemens Medical Systems, Erlangen, Germany) with the patient under general anaesthetic with ventilator parameters adjusted to keep end-tidal carbon dioxide between 3.5 - 5.5 kPa. Supplemental oxygen was given as required to maintain oxygen saturations (SpO_2_) at the usual pre-anaesthetic value for the patient.

#### Flow Imaging

Through-plane quantitative flow data was acquired using retrospectively gated, velocity encoded, phase contrast magnetic resonance. Images were either acquired using a free breathing Cartesian sequence with 3 signal averages or a spiral sequence acquired during a short apnoeic period of 5-8 seconds. ^5^ Data was acquired in the following positions: SVC close to pulmonary artery anastomosis, IVC at diaphragm level, pulmonary trunk (if present), proximal branch PAs, proximal pulmonary veins and ascending aorta. Vessels were segmented using a semi-automatic vessel edge detection algorithm (OsiriX; OsiriX Foundation, Switzerland) with manual operator correction. The following calculations was made using flow data: Systemic-to-pulmonary collateral flow proportion = (total pulmonary venous return - total PA flow) / total pulmonary venous return, expressed as a percentage.^6^

#### Ventricular Volume and Function

Ventricular volumes were assessed using a retrospectively gated multi-slice short-axis steady state free precession cine sequence. ^7^ Slices were acquired separately, in an apnoeic period of 5-10 seconds. Manual segmentation quantified end diastolic and systolic volumes (EDV and ESV) of the functionally single ventricle using an in-house plug-in for OsiriX. Stroke volume (SV) and ejection fraction (EF) were calculated from the volumetric data. Atrioventricular valve regurgitation (AVVR) was calculated from flow and volumetric data.

#### Anatomical assessment

Arterial and venous anatomy were assessed using gadolinium-enhanced MRA as previously described. ^8^ Two consecutive angiograms were acquired within a single 20-30 second period of apnoea. The first angiogram provided systemic arterial anatomy and the second angiogram provided second-pass contrast enhancement of venous and PA anatomy. Systemic venous decompressing collaterals from SVC territory to IVC territory were visualised using late-phase 3D MRA. These collaterals were graded by severity as previously described.^3^

### Measurement of central venous pressure during pre-operative CMR

Following CMR data acquisition, a right internal jugular venous line (Abbocath-T, 22G, Venisystems) was sited aseptically, under ultrasound guidance. ^9^ The mean central venous pressure (CVP_BCPC_) was transduced after careful flushing and zeroing, under the same conditions as the CMR, with interruption of positive pressure ventilation (passive expiration). Following measurement, the cannula was removed and the site dressed.

### Pressure-Flow Metrics

Pressure and flow data were used to calculate the following metrics (Figure 1):

**Figure 1.**
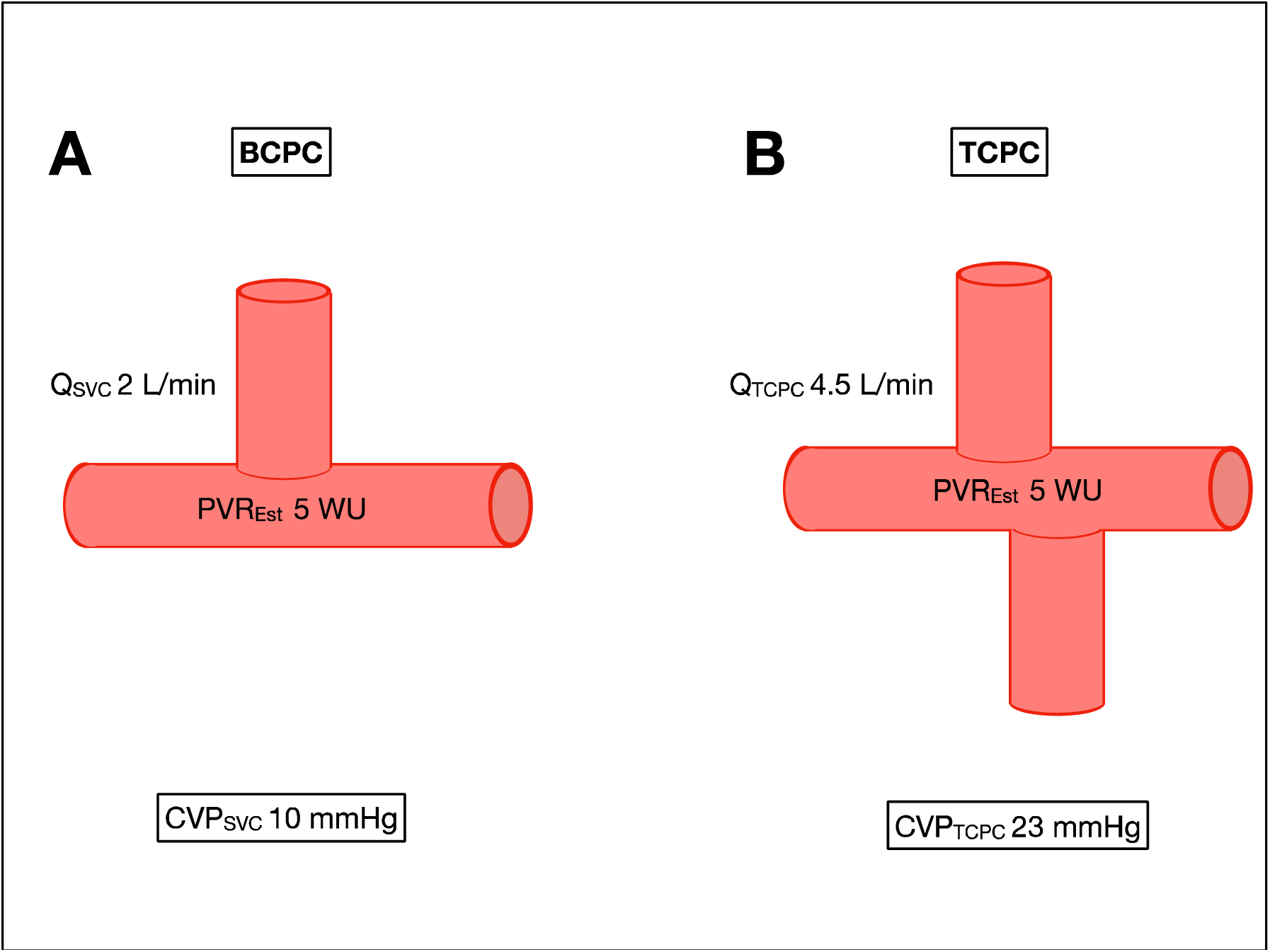
Diagrammatic presentation of methodology for calculating CVP_TCPC_. At BCPC stage, SVC flow and CVP_SVC_ are measured to calculate an estimate of PVR which neglects distal atrial pressure. B. The CVP_TCPC_ pressure is calculated using product of PVR and the assumed TCPC flow from either aortic flow or SVC + IVC flow.

1. A simple estimate of pulmonary vascular resistance (PVR_EST_) that neglects left atrial pressure, calculated by dividing CVP at time of BCPC by Q_p_ (SVC flow or SVC flow + native PA flow):

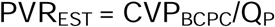
2. An estimate of CVP following completion of the TCPC (CVP_TCPC_) assuming post-TCPC pulmonary artery flow will equal aortic flow, Q_Ao_:

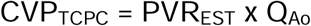

### Sensitivity Analysis to alternative method of measuring systemic flow

Estimated CVP_TCPC_ is calculated using aortic flow which necessarily includes systemic to pulmonary collateral flow (SPC). We also performed a sensitivity analysis using CVP_TCPC_ which excludes SPC flow (substituting Aorta flow with SVC+IVC flow).

### Predetermined outcome measures

Post-operative ICU electronic records were available for patients from 2012 onwards (n=70). In this group, the mean of hourly CVP in the 24hrs after TCPC (CVP_ICU_) was recorded for comparison to pre-operative CMR measures.

Early outcome was evaluated in two ways:

i. Length of hospital stay (LOS) (measured from the day of TCPC surgery until the day of discharge from hospital to home)
ii. Composite early outcome of need for emergency fenestration, emergency TCPC takedown or early death (<30 days post TCPC).

### Statistics

STATA 13.1 and Graphpad Prism 5f were used for statistical analysis and Figures. Data were examined for normality and where appropriate, non-normally distributed variables were log transformed to ensure normal distribution prior to analysis. Descriptive statistics are expressed as mean (±95% confidence interval) when normally distributed, and median (IQR) when non-normally distributed, unless specified. Proportions are expressed as percentages. Data were examined for normality using the Shapiro-Wilk test, and where appropriate, non-normally distributed variables were transformed prior to analysis.

We used logistic regression analysis to assess the relationship between EFF and clinical parameters. Multivariable logistic regression analysis was used to assess independent relationships (and control for confounding) between EFF and associated covariates. Covariates with a p<0.1 were eligible for inclusion in the multivariable model. The area under the receiver operating characteristics curve was used to identify the threshold of CVP_TCPC_ with the greatest classification accuracy (optimized sensitivity and specificity).^10^

### Patient Public Involvement

This research was done without patient involvement. Patients were not invited to comment on the study design and were not consulted to develop patient relevant outcomes or interpret the results.

## Results

### Demographics

CMR and central venous pressure (CVP_BCPC_) data were obtained in 131 patients (80 male) prior to TCPC completion under general anaesthesia. Patient characteristics for the study cohort are described in Table 1. There were no significant differences between the study cohort and the 16 excluded patients in terms of age, sex, cardiac morphology, cardiac output, ejection fraction, length of hospital stay or EFF.

**Table 1.**
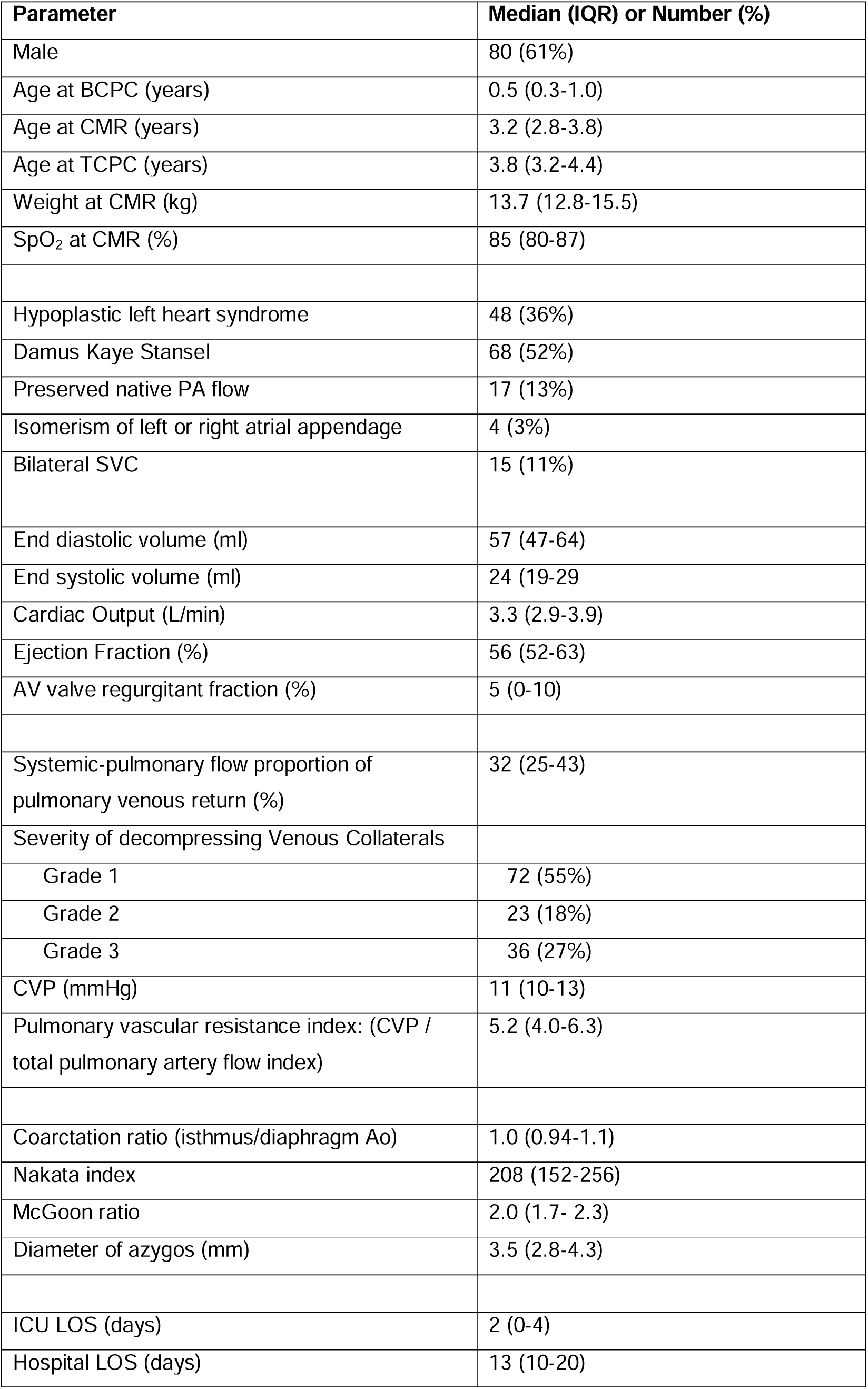
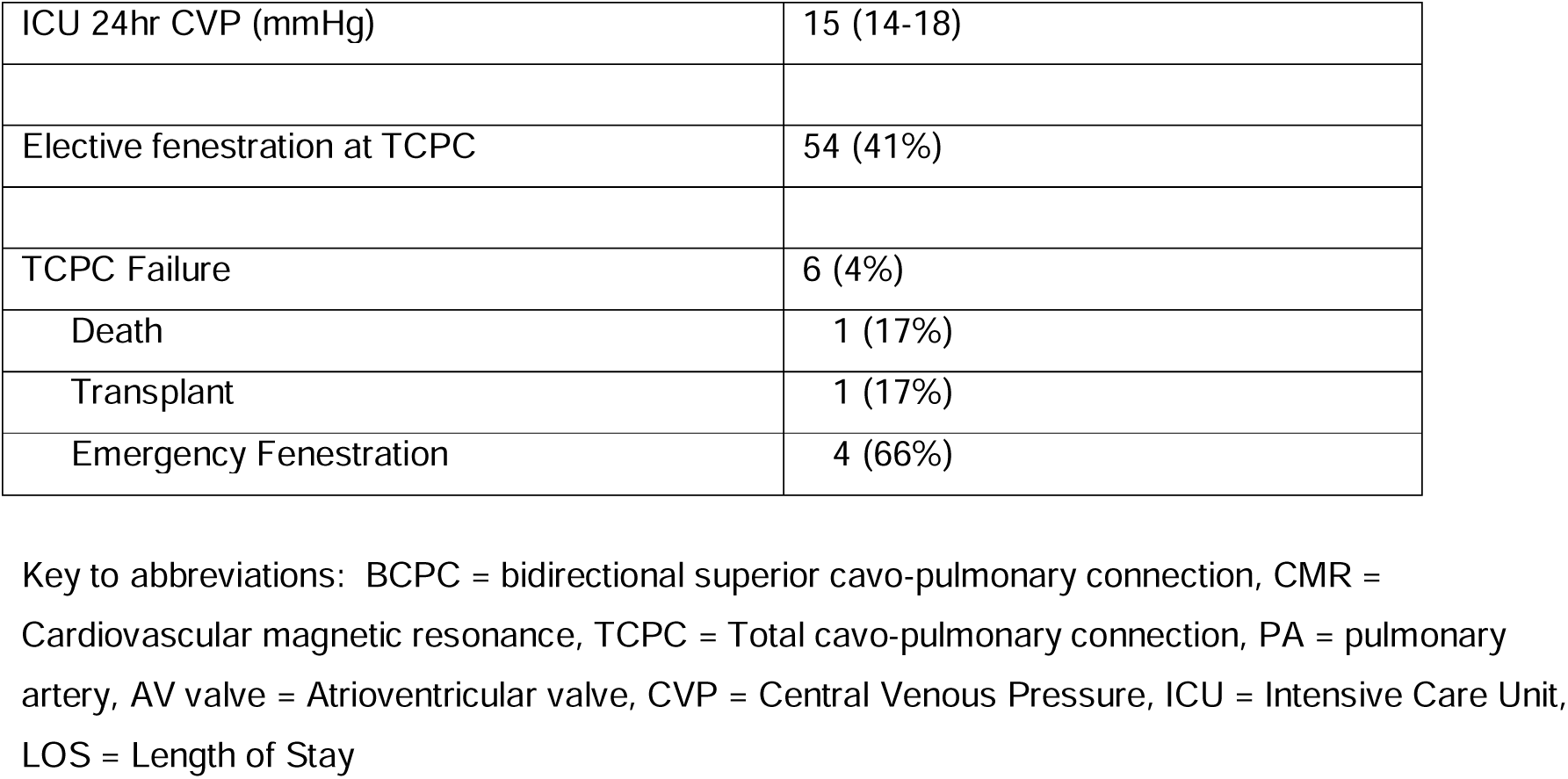
Patient demographics in the study cohort, n=131

The median age at CMR was 3.2years (IQR 2.8-3.8years) and age at TCPC completion 3.8 (IQR 3.2-4.4years). TCPC completion is performed in our institution using an extra-cardiac conduit and the TCPC was electively fenestrated in 41% of patients. Median CVP_TCPC_ was 23.6mmHg (IQR 18.1-28.4 [range 5.2-48]).

### Relationship to ICU Pressure

Post-operative ICU electronic records were available in 70 patients. Estimated CVP_TCPC_ correlated significantly with CVP_ICU_ (r=0.26, p=0.03), particularly in patients without a fenestration (n=33, r=0.45, p=0.01), Figure 2. However, CVP_TCPC_ significantly overestimated CVP_ICU_ (15±3 vs 22±7mmHg).

**Figure 2.**
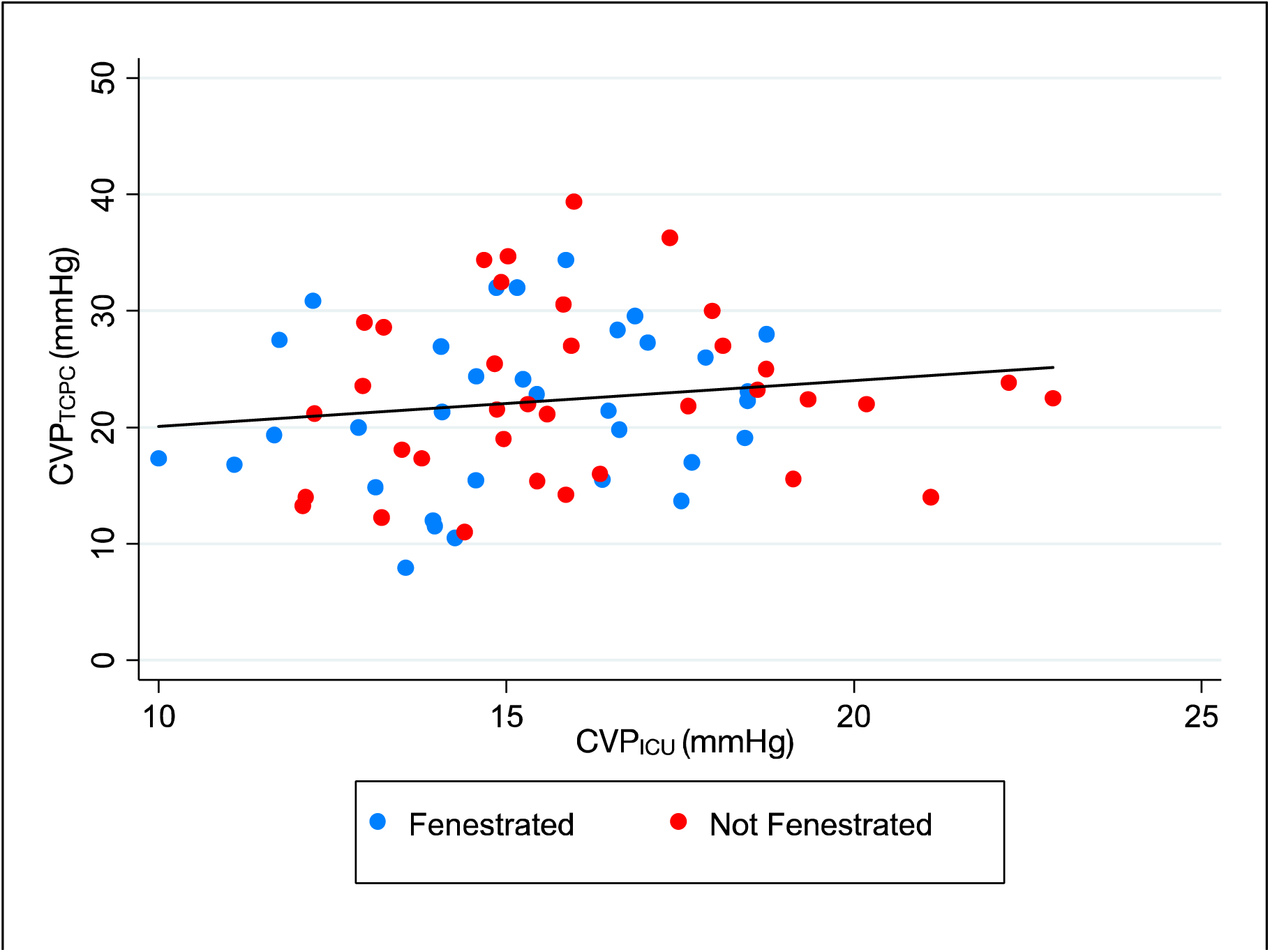
Scatter plot of CVP_TCPC_ and CVP_ICU_. Patients with fenestrated TCPC are shown in blue, compared with non-fenestrated in red.

### Relationship to clinical parameters

There was no association between CVP_TCPC_ and patient age at CMR, age at BCPC or sex. Patients with higher oxygen saturations at the time of CMR had lower estimated CVP_TCPC_ (Beta -0.19, p=0.047). CVP_TCPC_ was higher in patients with HLHS (27 vs 22mmHg, p<0.005), in whom there was a higher PVR_Est_ (6.1 vs 5.1WU index, p=0.01).

### Outcome

#### Hospital Stay

The duration of hospital stay was associated with estimated CVP_TCPC_ (Beta= -0.18, p=0.04). The duration of hospital stay was also associated with CVP_ICU_, hypoplastic left heart syndrome, the severity of offloading veno-venous collaterals, and the volume of systemic to pulmonary collaterals (Table 2). However, an independent association between hospital stay and estimated CVP_TCPC_ was not sustained in a multivariable linear regression analysis containing the other significant metrics, model r^2^ = 0.27 (Table 2).

**Table 2.** Univariable and multivariable linear regression associations between log-transformed hospital stay and exploratory variables.

| *Variable* | *Univariable* |  | *multivariable* |  |
| --- | --- | --- | --- | --- |
|  | *Beta* | *Significance* | *Beta* | *Significance* |
| CVP<sub>ICU</sub> | -0.32 | **0.008** |  |  |
| Estimated CVP<sub>TCPC</sub> | -0.18 | **0.04** | 0.12 | 0.9 |
| CVP<sub>BCPC</sub> | -0.01 | 0.9 |  |  |
| PVR<sub>EST</sub> | -0.11 | 0.2 |  |  |
| Severity of decompressing Venous Collaterals | -0.32 | **<0.005** | -0.39 | **<0.005** |
| SPC Flow | -0.25 | **0.006** | -0.24 | **0.01** |
| End-diastolic volume | 0.05 | 0.6 |  |  |
| Ejection Fraction | 0.004 | 1.0 |  |  |
| Hypoplastic Left Heart Syndrome | -0.28 | **<0.005** | -0.18 | **0.04** |

#### Early Fontan failure

EFF occurred in 6/131 patients during the study period: Emergency fenestration – 4, Emergency transplantation – 1, Death -1 (Table 1).

CVP_TCPC_ was significantly associated with EFF (Odds Ratio [OR] 1.12 (1.02-1.24), p=0.02). A threshold of CVP_TCPC_ ≥33mmHg was found to have the highest specificity (90%) and sensitivity (67%) for identifying EFF (area under receiver operating curve, AUC = 0.79), OR 18.8 (3.1-114), p=0.001, Figure *3*A.

**Figure 3.**
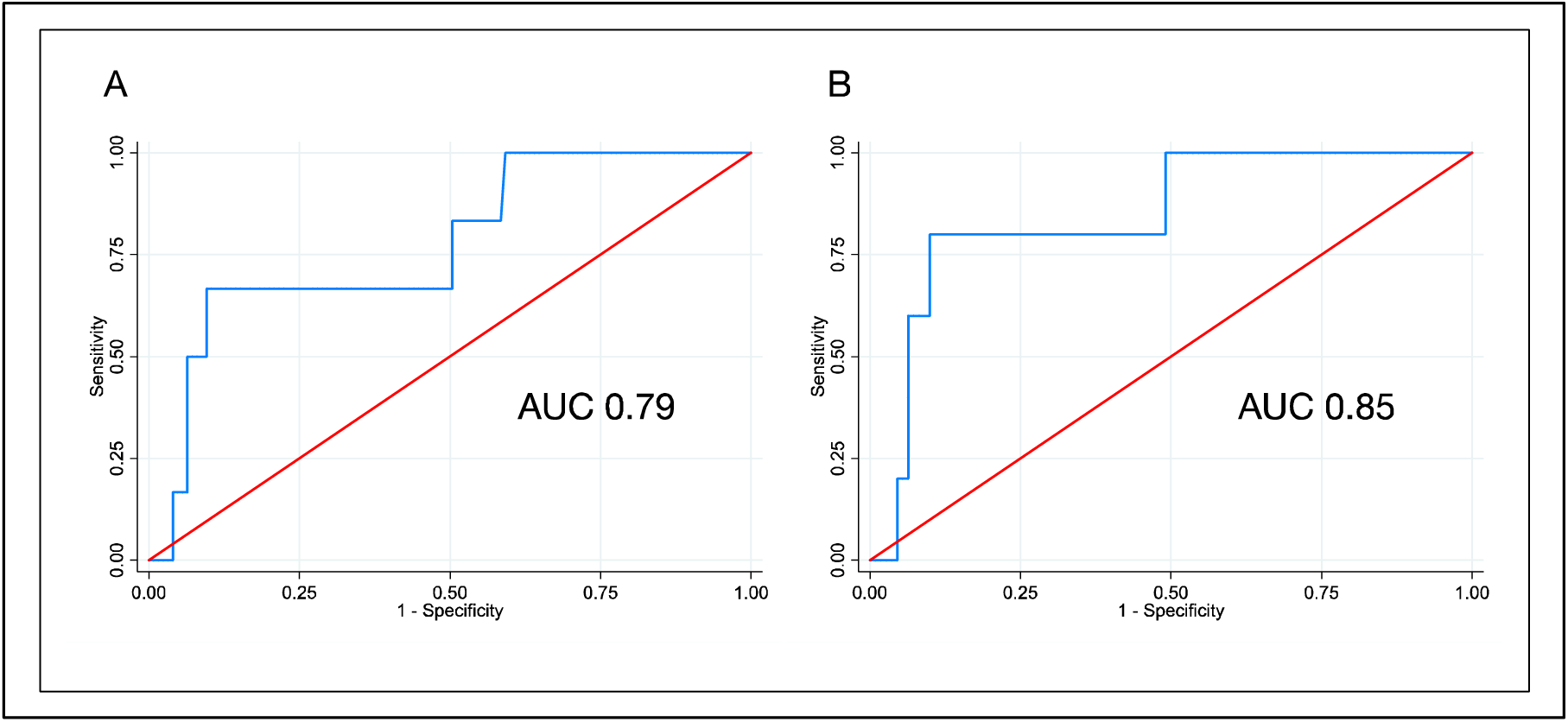
Receiver operating characteristic curves (ROC) for estimated CVP and early TCPC failure. A: All patients, AUC 0.79. Sensitivity 0.67 and specificity 0.90 at cut-point 33mmHg (OR 18.8, p=0.001). B: Patients with single SVC, AUC 0.85, Sensitivity 0.80 and specificity 0.90 at cut-point 33mmHg (OR 36, p=0.002).

The relationship between CVP_TCPC_ and EFF was stronger in patients with a single SVC (n=115, OR 1.2 [1.03-1.27], p=0.01). In this group, a CVP_TCPC_ threshold of ≥33mmHg was also found to have the highest specificity (90%) and sensitivity (80%) for EFF (AUC=0.85), OR 36.0 (3.7-351), p=0.002, Figure *3*B.

Except for the severity of systemic veno-venous collateral grade (p=0.046), there was no other univariable associations between EFF and conventional pre-operative CMR and demographic variables (including: CVP_BCPC_, ventricular volumes, ejection fraction, PVR_EST_, hypoplastic left heart syndrome, azygos vein diameter, SPC flow, age at TCPC, age at BCPC and sex), Figure 4.

**Figure 4.**
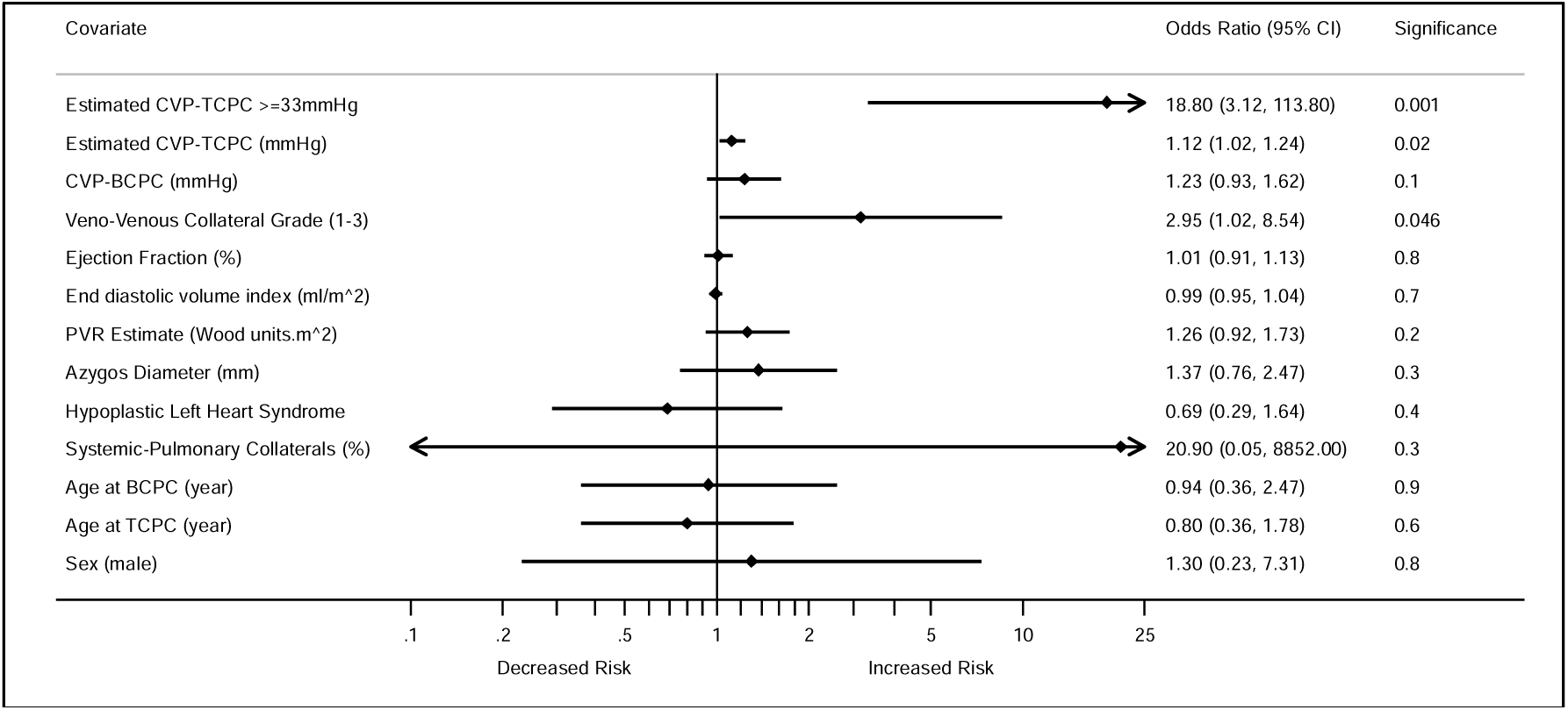
Forest plot of univariable associations between clinical covariates and EFF. Data is expressed as odds ratio (±95% confidence interval).

In a multivariable model containing systemic veno-venous collateral grade and CVP_TCPC_, only estimated CVP_TCPC_ predicted EFF (p=0.008).

### Sensitivity Analyses

#### Alternative method of measuring systemic flow

Estimated CVP_TCPC_ calculated by excluding SPC flow was significantly lower than with SPC flow included: 18 vs 24mmHg, p<0.05. Calculated in this manner, there remained an equally significant association with EFF (OR 1.2 (1.03-1.43), p=0.02). However, there was no significant correlation with CVP_ICU_ for the group (r=0.1, p=0.4) and only a trend to correlation in patients without fenestration (r=0.35, p=0.06)

## Discussion

With the evolution of surgical and perioperative management of the TCPC, biomarkers from previous eras may no longer prove robust. In this study we have shown that a novel estimated pressure metric, CVP_TCPC_, can be calculated from pre-operative data and that it is independently associated with early Fontan failure and is correlated with directly measured post-operative pressure from ICU.

Although EFF has decreased in incidence in published series, it is still an important clinical event.^4^ In this study we have used a conventional definition based on objective clinical events and investigated typical pre-operative risk factors. CVP_TCPC_ may perform well as a predictive biomarker in our series because it is closely related to the haemodynamic hallmark of the condition – high CVP.

Our analysis showed a reasonable correlation between measured CVP_ICU_ and estimated CVP_TCPC_. However, there was a significant bias of approximately 7mmHg. The most obvious explanation is that the post-operative haemodynamic state does not completely satisfy the assumptions of our calculation – these being: unchanged cardiac output, pulmonary vascular resistance and mean atrial pressure between the BCPC to TCPC. In fact, after the TCPC, increased systemic arterial saturation means that lower systemic flow is required to maintain oxygen delivery, while PVR may fall due to increased pulmonary flow. In addition, during the postoperative ICU period, therapeutic interventions (e.g. ventilation, inotropes, pulmonary vasodilators, fluids, diuretics, dialysis) are directed towards the modulation CVP. Thus, a significant correlation between CVP_TCPC_ and the directly measured pressure is slightly surprising.

The fact that CVP_TCPC_ is associated with EFF, even when its constituent components (Qs and PVR) don’t, suggest its importance as an integrator of deleterious haemodynamics. The stronger relationship between CVP_TCPC_ and clinical outcome in patients with single SVC is interesting and may be because accurate measurement of CVP_BCPC_ in patients with bilateral SVCs is more difficult due to asymmetric SVC size or pulmonary artery narrowing between the bilateral Glenn anastomoses. Nevertheless the diagnostic accuracy in the entire group remains satisfactory.

The clinical implications of these data are that it may be possible to identify patients at increased risk of EFF. Given further validation, such a metric could be used to improve selection of elective fenestration. Several institutions employ elective fenestration in all TCPCs, and the results of this study may not be directly translatable. There is currently a lack of consensus regarding routine fenestration; whilst it may reduce post-operative CVP, it comes at the expense of increased systemic desaturation and a possible increased risk of systemic thromboembolism.^11-14^

CVP_TCPC_ was not independently associated with hospital stay when included in a multivariable model with systemic to pulmonary collateral (SPC) flow, severity of venous collaterals and a diagnosis of HLHS. SPC flow has previously been shown to be strongly associated with hospital stay, operating through its association with prolonged pleural effusions.^6 15 16^ In our data CVP_TCPC_ and SPC flow are mathematically coupled, given the latter’s inclusion in the calculation of the former (in aortic flow). CVP_TCPC_ was also associated with a diagnosis of HLHS, being higher in patients with this diagnosis (due to higher PVR_Est_ in the HLHS patients). The lack of independence is likely related to collinearity with other variables. Assessment of CVP_TCPC_, systemic to pulmonary collaterals and veno-venous collaterals therefore may provide important and complementary data about the expected post-operative course.

## Limitations

This is a retrospective study from a single centre, which may limit generalisation of the study findings, insofar as our patient population and practice differ. However, our clinical practice will be broadly similar to many institutions. Nevertheless, one advantage of the retrospective design is that CVP_TCPC_ was not used during multidisciplinary meetings to guide decision making, and therefore will not have influenced clinical outcomes, such as the rate of EFF, decision to defer TCPC, or fenestration.

Our method of pre-operative clinical evaluation does not involve routine cardiac catheterization, therefore we are not able to evaluate the relationship of elevated end-diastolic pressure (independently of CVP) in our dataset.

## Conclusion

CVP_TCPC_ is easily calculated at the time of pre-TCPC assessment by combining pressure and flow data. Although there is a significant bias between estimated and measured CVP, higher CVP_TCPC_ is associated with an increased risk of EFF events. Thus, this metric could be used to inform important clinical decisions such as pre-emptive TCPC fenestration. However, further larger multi-centre prospective studies are required to validate this metric, especially in centres who undertake routine TCPC fenestration.

## Data Availability

Data is available upon reasonable request.

## Contributors

Each author has contributed significantly to the submitted work. MQ conceived the design of the study. The data collection, analysis and interpretation of the data was undertaken by MQ, IC, SS, MH and VM. The drafting of the manuscript and its revision was completed by MQ and VM. Each has read and approved the manuscript as written, and there are no conflicts of interest to disclose. None of the paper’s contents have been published previously and it is not under consideration elsewhere.

## Competing interests

None declared.

## Patient consent for publication

Not required.

## Ethics approval

This study was approved by the local research ethics committee.

## Provenance and peer review

Not commissioned; externally peer reviewed.

## Data availability statement

Data are available on reasonable request.

## Notes

### Competing Interest Statement

The authors have declared no competing interest.

### Funding Statement

No external funding was received for this work.

## References

1. Chaudhari M, Sturman J, O’Sullivan J, et al. Rescue cardiac transplantation for early failure of the Fontan-type circulation in children. The Journal of Thoracic and Cardiovascular Surgery 2005;129(2):416–22. doi: https://doi.org/10.1016/j.jtcvs.2004.06.030

2. Ohuchi H, Miyazaki A, Negishi J, et al. Hemodynamic determinants of mortality after Fontan operation. Am Heart J 2017;189:9–18. doi: 10.1016/j.ahj.2017.03.020 [published Online First: 2017/06/20]

3. Hughes ML, Broadhead M, McEwan A, et al. Pre-Operative Grade of Decompressing Systemic Venous Collaterals, But Not Jugular Venous Pressure, Predicts Short-and Medium-Term Outcome After Completion of the Total Cavopulmonary Connection. JACC: Cardiovascular Imaging 2019:2903. doi: 10.1016/j.jcmg.2018.12.017

4. Murphy MO, Glatz AC, Goldberg DJ, et al. Management of early Fontan failure: a single-institution experience. European Journal of Cardio-Thoracic Surgery 2014;46(3):458–64.

5. Steeden JA, Atkinson D, Hansen MS, et al. Rapid flow assessment of congenital heart disease with high-spatiotemporal-resolution gated spiral phase-contrast MR imaging. Radiology 2011;260(1):79–87. doi: 10.1148/radiol.11101844 [published Online First: 2011/03/19]

6. Odenwald T, Quail MA, Giardini A, et al. Systemic to pulmonary collateral blood flow influences early outcomes following the total cavopulmonary connection. Heart 2012;98(12):934–40. doi: 10.1136/heartjnl-2011-301599 [published Online First: 2012/05/26]

7. Quail MA, Nordmeyer J, Schievano S, et al. Use of cardiovascular magnetic resonance imaging for TAVR assessment in patients with bioprosthetic aortic valves: comparison with computed tomography. Eur J Radiol 2012;81(12):3912–7. doi: 10.1016/j.ejrad.2012.07.014 [published Online First: 2012/08/15]

8. Hughes ML, Tsang VT, Kostolny M, et al. Lessons from inter-stage cardiac magnetic resonance imaging in predicting survival for patients with hypoplastic left heart syndrome. Cardiol Young 2011;21(6):646–53. doi: 10.1017/S104795111100059X

9. Stockton E, Hughes M, Broadhead M, et al. A prospective audit of safety issues associated with general anesthesia for pediatric cardiac magnetic resonance imaging. Paediatr Anaesth 2012;22(11):1087–93. doi: 10.1111/j.1460-9592.2012.03833.x

10. Liu X. Classification accuracy and cut point selection. Statistics in medicine 2012;31(23):2676–86.

11. Bridges ND, Lock JE, Castaneda AR. Baffle fenestration with subsequent transcatheter closure. Modification of the Fontan operation for patients at increased risk. Circulation 1990;82(5):1681–9. doi: 10.1161/01.cir.82.5.1681 [published Online First: 1990/11/01]

12. Hsu DT, Quaegebeur JM, Ing FF, et al. Outcome after the single-stage, nonfenestrated Fontan procedure. Circulation 1997;96(9 Suppl):Ii-335-40. [published Online First: 1997/12/31]

13. Thompson LD, Petrossian E, McElhinney DB, et al. Is it necessary to routinely fenestrate an extracardiac fontan? J Am Coll Cardiol 1999;34(2):539–44. doi: 10.1016/s0735-1097(99)00228-4 [published Online First: 1999/08/10]

14. Lemler MS, Scott WA, Leonard SR, et al. Fenestration improves clinical outcome of the fontan procedure: a prospective, randomized study. Circulation 2002;105(2):207–12. doi: 10.1161/hc0202.102237 [published Online First: 2002/01/16]

15. Glatz AC, Rome JJ, Small AJ, et al. Systemic-to-pulmonary collateral flow, as measured by cardiac magnetic resonance imaging, is associated with acute post-Fontan clinical outcomes. Circ Cardiovasc Imaging 2012;5(2):218–25. doi: 10.1161/CIRCIMAGING.111.966986 [published Online First: 2012/01/10]

16. Grosse-Wortmann L, Drolet C, Dragulescu A, et al. Aortopulmonary collateral flow volume affects early postoperative outcome after Fontan completion: A multimodality study. J Thorac Cardiovasc Surg 2012 doi: 10.1016/j.jtcvs.2012.03.032 [published Online First: 2012/04/17]

